# Autoimmune and inflammatory comorbidity patterns in rheumatoid arthritis: temporal trajectories and impact on persistence on DMARD therapy

**DOI:** 10.1101/2025.05.04.25326920

**Authors:** Signe Hässler, Julie Aste, Francis Berenbaum, Michelle Rosenzwajg, Jérémie Sellam, David Klatzmann, Milka Maravic

**Affiliations:** Biotherapy (CIC-BTi) and Inflammation, Immunopathology, Biotherapy Department (i2B), Pitié-Salpêtrière Hospital, Assistance Publique-Hôpitaux de Paris, Paris, France; IQVIA Operations France, General management, Courbevoie, Paris, France; IQVIA Operations France, Data & IA, Courbevoie, Paris, France; INSERM UMRS 938, Centre de Recherche Saint-Antoine, FHU PaCeMM, Sorbonne Université, and Rheumatology Department, Saint-Antoine Hospital, Assistance Publique-Hôpitaux de Paris, Paris, France; Immunology, Immunopathology, Immunotherapy (i3), Sorbonne Université, INSERM, Paris, France; Rheumatology Department, Lariboisière Hospital, Assistance Publique-Hôpitaux de Paris, Paris, France

## Abstract

**Objective:** Patients with immunological diseases exhibit distinct comorbidity patterns, categorized into low comorbidity, polyautoimmunity, and polyinflammation clusters. This retrospective cohort study aimed to validate these profiles in rheumatoid arthritis (RA), examine their longitudinal trajectories, and assess their impact on treatment persistence.

**Methods:** RA patients receiving targeted therapies, and their associated treatments, were identified from the French pharmacy dispensing database LRx. Comorbidity clusters were assigned using a multinomial regression model, and state sequence analysis with hierarchical clustering was used to define temporal trajectories. Cox regression models evaluated DMARD persistence across trajectories.

**Results:** Among 15,189 RA patients (maximum follow-up: 10 years), initial comorbidity profiles included low comorbidity (61.9%), polyautoimmunity (24.7%), and polyinflammation (13.4%). Four trajectory patterns emerged: stable low comorbidity (50%), dominant polyautoimmune (31%), stable polyinflammatory (9%), and low comorbidity switching to polyinflammation (polyinflammation switchers, 10%). The prevalence of polyautoimmunity and polyinflammation increased with age by 2.5% and 3.8% per decade, respectively.

Patients with stable polyinflammation had the lowest classical synthetic DMARD persistence (HR: 1.79 [1.33–2.42], reference: polyinflammation switchers). Stable low comorbidity patients had the highest biological and targeted synthetic DMARD persistence (polyinflammation switchers aHR: 1.32 [1.09–1.60], reference: stable low comorbidity).

**Conclusion:** Comorbidity trajectories in RA influence DMARD persistence, reflecting distinct etiopathological subgroups with potential theranostic relevance.

## Introduction

Patients with rheumatoid arthritis (RA) are characterized by a high burden of multimorbidity, which is raising an increasing attention for its possible implications in patient care^1,2^. Defined as the coexistence of at least two long-term conditions in a patient without considering a “privileged” index disease, multimorbidity is present in 40-60% of RA patients and is more prevalent in RA than in the general population^3,4^. The related term comorbidity refers to the coexistence of a condition with an index disease but is often used as a synonym of multimorbidity.^1,2,4^

Studies in the last 20 years have reported an association of multimorbidity in RA with a lower quality of life and physical function, higher disease activity, mortality and difficulty to achieve disease remission.^5–10^ DMARD efficacy and retention is decreased in multimorbid RA and specific comorbidities have been associated with some adverse events on therapy.^11–16^

Since the diseases included in multimorbidity may vary from one study to another, multimorbidity indexes counting pre-defined lists of conditions have been developed to harmonize the methodology.^17^ These indexes are appropriate for adjusting statistical models for the confounding due to comorbidities, but they do not take into account the correlations between comorbidities.^18^ To date only a few studies have performed unsupervised analyses to identify multimorbidity patterns in RA and they have consistently identified a cardiopulmonary, cardiometabolic and chronic pain/mental health patterns.^19,20^

Despite these recent advancements in the study of multimorbidity, a critical question remains the choice of the diseases to be included, which ranges between 11-50 in different studies and is restricted to concomitant long-term conditions. While this is highly relevant for patient care, it might not necessarily inform on etiopathogenic heterogeneity within RA, where other minor non-severe comorbidities and even past conditions might be informative. In an effort to identify etiopathogenic clusters we recently performed an unsupervised analysis on 375 patients with one of 19 autoimmune and inflammatory diseases (AIDs) using the 30 most prevalent past and present comorbidities.^21^ Since one of the aims was to improve and personalize patient classification within the immunological disease continuum,^22^ among the 30 comorbidity variables we included the number of autoimmune and of inflammatory diseases, which are never considered in multimorbidity studies. We described 3 mutually exclusive comorbidity profiles characterized by: i) low comorbidity burden; ii) polyautoimmunity, allergy, history of viral infections; iii) polyinflammation, dyslipidemia, hypertension, obesity, mood disorders, cancer. RA patients were distributed among the 3 patterns and presented different clinical characteristics according to their comorbidity profile, with higher DAS28 and more extrarticular manifestations in the polyautoimmunity and polyinflammation profiles, higher pain and lower ACPA and RF seropositivity in the polyinflammation profile.^21^

To assess the validity of these profiles for patient stratification here we apply them to RA patients from a retail-pharmacy drug dispensation database and we evaluate their temporal stability during a 10-year follow-up and their association with DMARD therapy retention. We hypothesized that comorbidity profiles would be stable or evolve from low comorbidity to either polyautoimmunity or polyinflammation, and that persistence on DMARD therapy would be different among comorbidity profiles.

## Patients and Methods

### Study population

In this study we analyze RA patients and controls from LRx (Lifelink Treatment Dynamics), an IQVIA database containing anonymized outpatient care drug dispensing data from a panel of 10,000 pharmacies.^23^ In France RA is considered a long-term condition and RA therapies are 100% reimbursed by the healthcare system. To identify RA patients, any patient who received an oral or subcutaneous targeted therapy (TT) for a chronic inflammatory disease in 2023 was selected (raw patients: 269084). The disease leading to the prescription of a TT was identified using an algorithm based on the date of the TT indication, the specialty of the prescriber, and specific medication tracking baskets for certain diseases (BTSD algorithm developed by IQVIA).^24–26^ Details of the BTSD algorithm used to identify RA patients are described in the supplementary methods. We numbered a raw number of 46143 RA patients (Figure 1).

**Figure 1.**
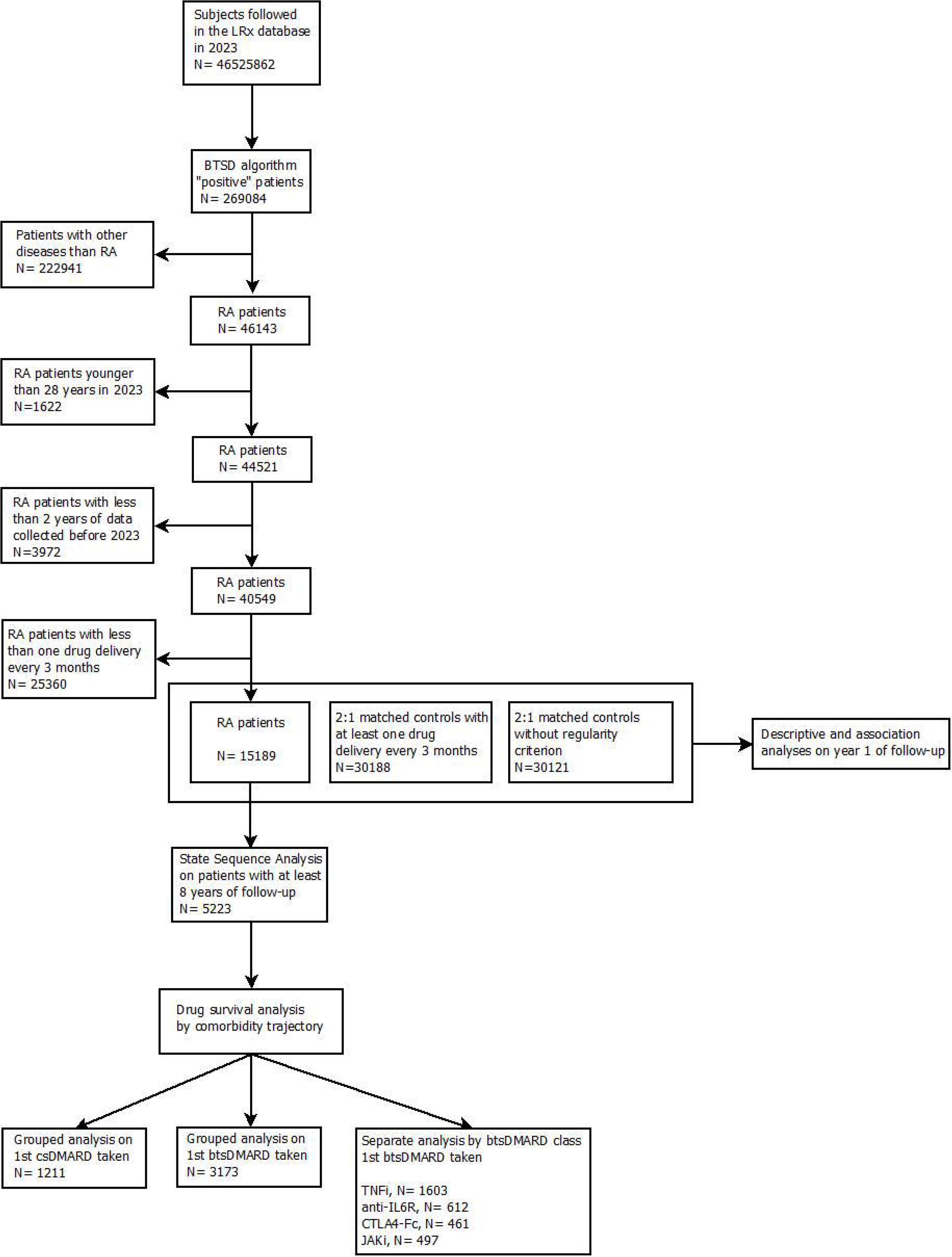
Study flow chart

RA patients were eligible if they were 28 years or older in 2023 (and therefore at least 18 years in 2014), if they had at least two years of drug delivery data collected and a regularity criterion defined as at least one drug delivery every 3 months. Age, sex and all the drug delivery data including drug name, European Pharmaceutical Market Research Association (EPhMRA) code, number of units of delivered drug and date of drug delivery from 01-01-2014 until 31-12-2023 were extracted and used for the state sequence and drug survival analyses (cohort study, Figure 1 and Supplementary Figure 1A and C). According to General Data Protection Regulation (GDPR), a maximum of 10 years of data is used per patient^27^.

Controls were identified as never having taken any biological nor targeted synthetic DMARD (btsDMARD) during the follow-up. They were eligible if they were 18 years or older on their first year of follow-up. Controls were matched in a 2:1 proportion to RA patients on sex, age classes ([19-29), [30-49), [50-69), [70-97)), geographical region of attended pharmacy and year of start of follow-up. Two control populations were selected, one with the same regularity criterion as RA and one without. Only the drug delivery data of the first two years of follow-up was extracted for the controls. All the RA patients and controls were included in the descriptive and association analyses (case-control study, Figure 1, Supplementary Figure 1B).

Data processing was performed in compliance with applicable laws including GDPR. The LRx database was authorized by the French Data Protection Authority (CNIL) on the 21st of October 2011 [reference: DE-2011-097] and updated on July 2018 for compliance with the GDPR [reference: DE-2018-289].

This study complies with the guidelines for Reporting of Studies Conducted Using Observational Routinely Collected Data for Pharmacoepidemiology (RECORD-PE).^29^

### Study variables

Three comorbidity profiles were previously identified through an unsupervised analysis on patients and controls from the TRANSIMMUNOM study^21,28^ using 30 comorbidity variables.^21^ Only 17 out of the 30 variables originally used could be defined by usage of specific drugs in LRx. We therefore designed algorithms to define 17 comorbidity variables from drug usage, to be able to apply on LRx a classification model of comorbidity clusters built on TRANSIMMUNOM patients. Anxiety, depression, dyslipidemia, hypertension, and respiratory tract disorder were defined using previously published algorithms.^30^ Ten other non-immunological comorbidities were built from knowledge of drug indications in the literature and in the French drug dictionary.^31^ A detailed description of the algorithms is given in the supplementary methods. Each algorithm was applied once per year using all the drug deliveries of the year starting from the first until the 10^th^ year of follow-up, thus generating up to 10 longitudinal disease variables per patient per algorithm (Supplementary Figure 1A).

To classify RA patients and controls from LRx into comorbidity clusters we developed a classification model on the TRANSIMMUNOM study. Details on the classification model are provided in the supplementary methods.

Demographic characteristics, DMARD and non-DMARD drug usage during the first year of follow-up of RA patients was compared among comorbidity profiles through one-way ANOVA for continuous variables and Fisher’s exact test for qualitative variables. The significance level for all the analyses was set at p<0.05 with two-sided tests.

### State Sequence Analysis

To investigate the stability of comorbidity profiles, temporal sequences of yearly comorbidity profiles during 10 years were analyzed through State Sequence Analysis (SSA). Patient sequences were left-aligned at database entry and only sequences with at least 80% completeness (i.e. at least 8 years available) were eligible for the SSA (Figure 1). The alternative states of the sequences were low comorbidity, polyautoimmunity and polyinflammation. Sequences were compared two-by two through optimal matching (OM), a dissimilarity measure considering insertions, deletions and substitutions necessary to make two sequences similar. An equal cost was attributed to insertions, deletions and substitutions and the OM distance was calculated as the minimal cost necessary to convert a sequence into another. To identify the most common homogeneous temporal trajectories of comorbidity profiles, agglomerative hierarchical clustering with Ward’s method was applied to the OM distance matrix. The number of trajectory clusters was selected through visual inspection of the dendrogram and of the sequence index plots.

To investigate the correlation between age and the proportion of RA patients or controls belonging to the different comorbidity profiles during the first year of follow-up, we estimated Pearson’s correlation after dividing age at year 1 in 5 years ordinal classes and smoothing the age of each patient to the minimum age of the class to which he belonged. For each comorbidity cluster proportion we performed a linear regression with age to estimate the rate of switch from low comorbidity to the other profiles.

### Drug survival analysis

To compare persistence on DMARD therapy among cluster trajectories a time to event analysis was performed. The outcome of interest was composite of two possible events, namely: 1) washout of the DMARD without renewal delivery; 2) delivery of a different DMARD, covering both the case of add-on therapy or of therapy switch to a different therapeutic class (Supplementary Table 5). Whichever of these came first during the first 18 months from DMARD initiation was considered as the event. The period covered by each drug delivery was estimated using package size, unit dose and usually prescribed daily dose, and the washout of the DMARD was considered as 3 times the period covered by the drug delivery (Supplementary Figure 1C). Persistence at 18 months from DMARD initiation was estimated through the Kaplan-Meier method.

Concomitant treatments were defined as at least 3 drug deliveries (glucocorticoids) or 1 drug delivery (other DMARDs) during the 3 months before and/or the 3 months after DMARD initiation (Supplementary Table 5). Previous treatments were defined as treatments whose last delivery was given any time within 10 years before DMARD initiation.

A global analysis on csDMARDs and btsDMARDs and a separate analysis by btsDMARD therapeutic class were performed (Figure 1, Supplementary Table 5). In the global analysis, the date of the first delivery of the csDMARD/btsDMARD was taken as index date and patients were eligible if they had no delivery of the same csDMARD/btsDMARD during the 12 months prior to the DMARD start; if more than one class of csDMARD or btsDMARD was taken during the follow-up only the first class taken was considered. For csDMARDs analysis no btsDMARD delivery was allowed before or at the date of csDMARD start (Figure 1, Supplementary Figure 1C).

Univariate analyses of drug survival by trajectory clusters, sex, age and concomitant treatments were done through log-rank tests. Multivariate Cox regression models were stratified on year of DMARD start and included trajectory clusters when univariate tests were significant at p<0.05 and other adjustment covariates when their univariate p<0.2. Only a minority of patients had missing data on sex (<0.8%), in which case multivariate models were performed on complete cases.

All the analyses were performed with R statistical software version 4.1.1. Additional packages used were caret 6.0-94, glmnet 4.1-7, gbm 4.2.3 and cvms 1.6.0 for classification model development, TraMineR 2.2-7 and fastcluster 1.2.3 for SSA, survival 3.2-11 and survminer 0.4.9 for drug survival analysis.

## Results

### Demographics and comorbidity profiles of RA patients in LRx

Through the BTSD algorithm and after removing patients with other exclusion criteria we identified 15189 RA patients in LRx eligible for analysis (Figure 1). The mean age at start of follow-up was 57.6 years and 64.5 % were women.

When comparing the demographic characteristics between comorbidity profiles the polyinflammation cluster had a significantly higher age while the polyautoimmunity cluster had a higher proportion of women compared to the other clusters (Table 1), and this was consistent to what previously observed in RA patients of the TRANSIMMUNOM study (Supplementary Table 8). Consistency was also seen for the proportion of RA patients in the clusters, which was highest in the low comorbidity and smallest in the polyinflammation cluster in both TRANSIMMUNOM and LRx (Supplementary Table 8). The most frequent therapy in the first year of follow-up were btsDMARDs alone and they were taken more frequently in the low comorbidity cluster, while patients in the polyautoimmunity and polyinflammation clusters took more oral glucocorticoids alone (Table 1). Antipyretics, proton pump inhibitors and folic acid were the most common concomitant medications (Supplementary Table 9). Apart from NSAIDs all the top 10 medications were taken more by patients in the polyautoimmunity and polyinflammation than in the low comorbidity cluster (Supplementary Table 9).

**Table 1.** Demographic characteristics and therapy of RA patients during the first year of follow-up in LRx.

|  | Low comorbidity<br>(N=9403) | Polyautoimmunity<br>(N=3745) | Polyinflammation<br>(N=2041) | p-value |
| --- | --- | --- | --- | --- |
| **Age (years)** |  |  |  |  |
| Mean (SD) | 55.9 (12.0) | 59.6 (11.7) | 62.1 (10.9) | <0.001 |
| Median [Min, Max] | 56.0 [19.0, 94.0] | 60.0 [19.0, 91.0] | 63.0 [24.0, 97.0] |  |
| **Sex** |  |  |  |  |
| Female | 5671 (60.3%) | 2807 (75.0%) | 1270 (62.2%) | <0.001 |
| Male | 3687 (39.2%) | 909 (24.3%) | 763 (37.4%) |  |
| Missing | 45 (0.5%) | 29 (0.8%) | 8 (0.4%) |  |
| **Follow-up (years)** |  |  |  |  |
| Mean (SD) | 5.83 (2.91) | 5.93 (2.99) | 6.13 (3.05) | <0.001 |
| Median [Min, Max] | 5.00 [2.00, 10.0] | 5.00 [2.00, 10.0] | 5.00 [2.00, 10.0] |  |
| **btsDMARD only** |  |  |  |  |
| Yes | 3478 (37.0%) | 1198 (32.0%) | 654 (32.0%) | <0.001 |
| No | 5925 (63.0%) | 2547 (68.0%) | 1387 (68.0%) |  |
| **csDMARD only** |  |  |  |  |
| Yes | 1579 (16.8%) | 597 (15.9%) | 317 (15.5%) | 0.253 |
| No | 7824 (83.2%) | 3148 (84.1%) | 1724 (84.5%) |  |
| **no DMARD** |  |  |  |  |
| Yes | 784 (8.3%) | 397 (10.6%) | 237 (11.6%) | <0.001 |
| No | 8619 (91.7%) | 3348 (89.4%) | 1804 (88.4%) |  |
| **csDMARD and btsDMARD association** |  |  |  |  |
| Yes | 974 (10.4%) | 363 (9.7%) | 168 (8.2%) | 0.0125 |
| No | 8429 (89.6%) | 3382 (90.3%) | 1873 (91.8%) |  |
| **Oral glucocorticoid only** |  |  |  |  |
| Yes | 612 (6.5%) | 328 (8.8%) | 176 (8.6%) | <0.001 |
| No | 8791 (93.5%) | 3417 (91.2%) | 1865 (91.4%) |  |
| **csDMARD and oral glucocorticoid association** |  |  |  |  |
| Yes | 664 (7.1%) | 268 (7.2%) | 170 (8.3%) | 0.13 |
| No | 8739 (92.9%) | 3477 (92.8%) | 1871 (91.7%) |  |

### Temporal trajectories of comorbidity profiles

To identify the most common temporal trajectories during 10 years of follow-up we performed a SSA using the comorbidity profiles as alternative states. After hierarchical clustering of the dissimilarity matrix we identified 4 clusters of temporal patterns (Figure 2 and Supplementary Figure 4). Three patterns were characterized by the dominance of one profile (patterns 1, 2 and 3 in Figure 2 and Supplementary Figure 4), and for two of them the dominant profile was relatively stable (pattern 2 with stable polyinflammation and pattern 3 with stable low comorbidity, Figure 2 and Supplementary Figure 4). Only pattern 4 was characterized by a progressive switch from low comorbidity to polyinflammation (Figure 2 and Supplementary Figure 4), but this pattern was present in only 544 out of 5223 sequences (10.4%).

**Figure 2.**
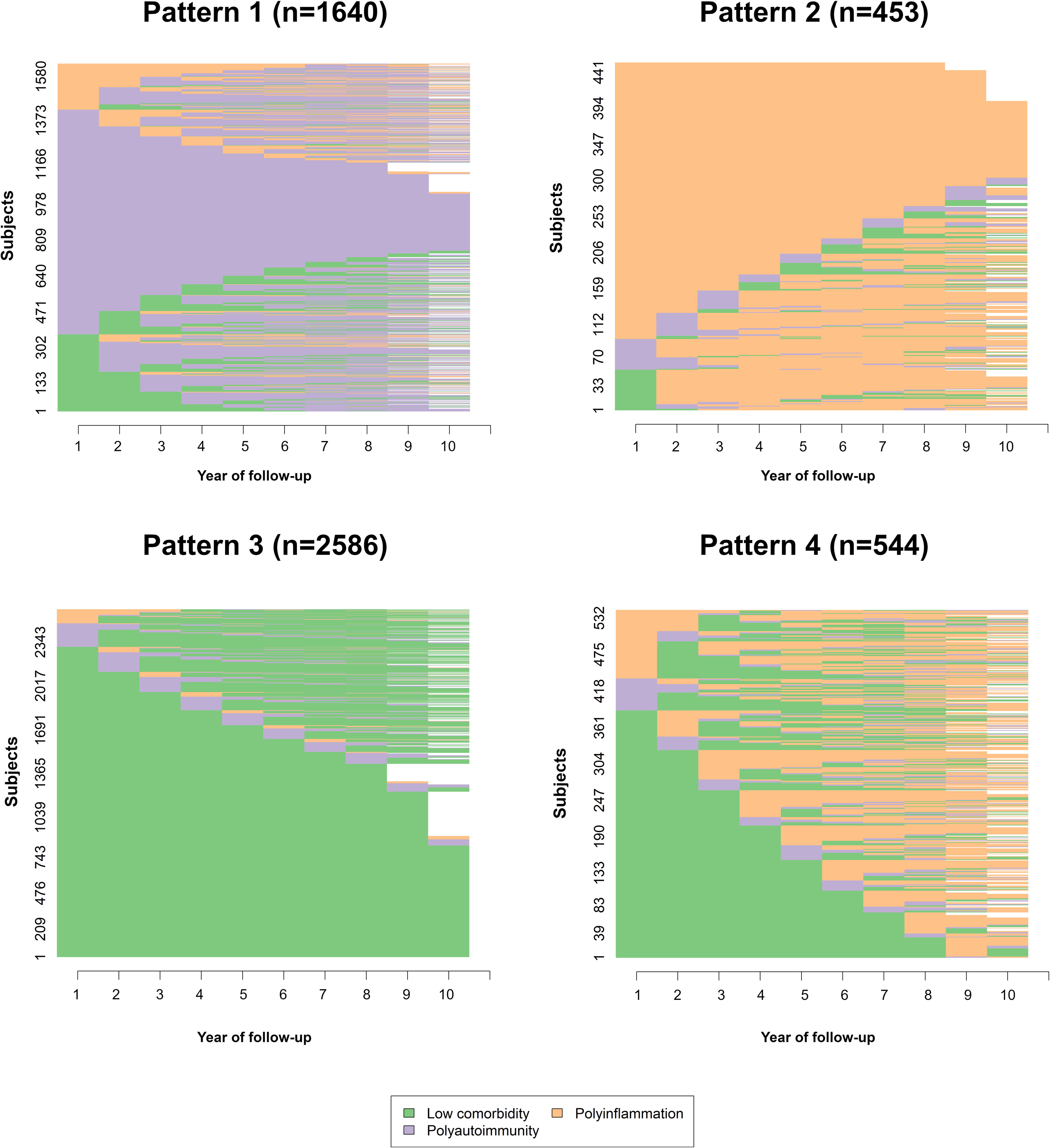
State sequence analysis sequence index plots of identified clusters of sequences with similar temporal trajectory.

The limited switch between comorbidity profiles observed during 10 years might be due to the fact that comorbidities are developed during aging and a longer period of observation is needed. Therefore we analyzed the proportion of RA patients belonging to the comorbidity profiles by 5-year age classes at the first year of follow-up (Figure 3A). There was a strong negative correlation between age and proportion of RA patients with low comorbidity and a positive correlation with the proportion in polyautoimmunity and polyinflammation (Figure 3A, Supplementary Table 10). The linear regression coefficients suggested that both the proportion with polyautoimmunity and with polyinflammation increased at a slow rate of 2.5 % and 3.8 % every 10 years respectively (Figure 3A, Supplementary Table 10). The proportion of patients with polyautoimmunity at 20 years of age was 14.8% on average, while it was only 0.1% for polyinflammation (Figure 3A).

**Figure 3.**
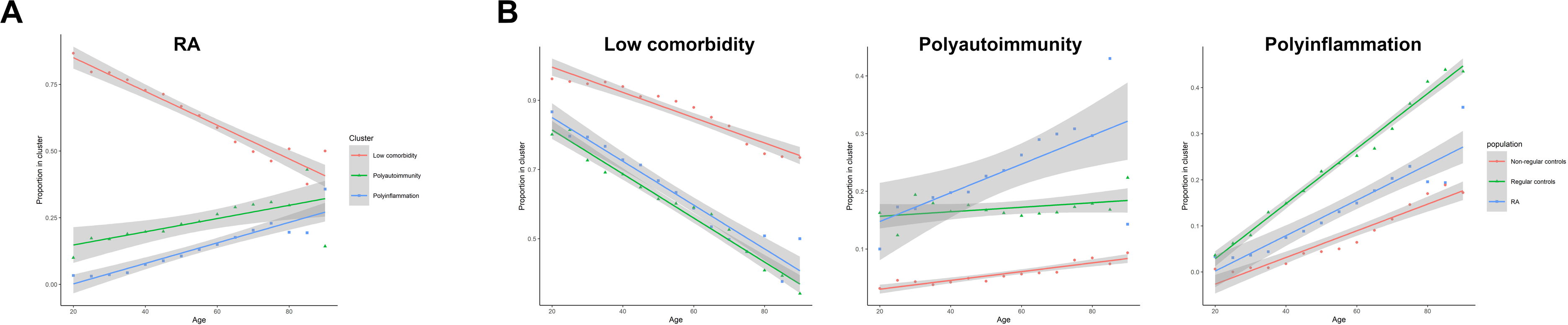
Scatter plot and linear regression of proportion of patients in comorbidity profiles by 5-year smoothed age at year 1 of follow-up in LRx. (A) RA patients; (B) Comparison of RA patients with regular and non-regular controls by comorbidity profile: low comorbidity (left panel), polyautoimmunity (middle panel), polyinflammation (right panel).

Since not only RA patients but also the general population develop comorbidities with aging we decided to compare RA with two matched control populations, one with the same dispensation regularity criterion as RA (regular controls, Figure 1, Supplementary Figure 1B and Supplementary Table 10), representative of patients with other diseases than RA, and the other without regularity criterion (non-regular controls, Figure 1, Supplementary Figure 1B and Supplementary Table 10), representative of the general population. The same algorithms and classification model previously developed for RA were applied to the controls followed by linear regression of the proportion of patients in comorbidity profiles by 5-year smoothed age. A larger proportion of non-regular than regular controls and RA were in the low comorbidity profile (p<0.001, Supplementary Table 11), while regular controls had more patients in the polyinflammation and less in the polyautoimmunity profile compared to RA (Supplementary Table 11). The linear regression revealed a more rapid decrease in the proportion of low comorbidity in RA patients and regular controls than in the non-regular controls (Figure 3B, left panel and Supplementary Table 10). The proportion of patients in polyautoimmunity increased significantly with aging in RA but not in regular controls, while it increased at a 3 times slower rate for non-regular controls than RA (Figure 3B, middle panel and Supplementary Table 10). All the populations started with similarly low proportions of patients with polyinflammation at 20 years, but they increased at significantly different rates with aging, with the slowest rate for non-regular controls, the highest for regular controls and an intermediate rate for RA (Figure 3B, right panel and Supplementary Table 10).

### Comparison of DMARD persistence between comorbidity trajectories

To compare therapy efficacy and drug tolerance between comorbidity trajectories we performed a drug survival analysis on csDMARDs and btsDMARDs. At the first initiation of a csDMARD, RA patients with stable polyinflammation and with dominant polyautoimmunity had a significant decrease in persistence compared to polyinflammation switchers (HR 1.79 [1.33; 2.42] and 1.33 [1.04; 1.71] respectively), while neither age, sex, previous or concomitant oral glucocorticoids were associated with csDMARD persistence in univariate analysis (Figure 4A). When initiating a btsDMARD, patients with stable low comorbidity had a significantly higher persistence compared to all the other trajectories (p=0.035, Figure 4B). Other factors associated with btsDMARD persistence in univariate analysis were age, sex, concomitant csDMARDs and glucocorticoids and previous glucocorticoids. In multivariable Cox regression models the different persistence between comorbidity trajectories was confirmed, with the lowest persistence for polyinflammation switchers compared to stable low comorbidity (HR 1.32 [1.09-1.6], Supplementary Figure 5A). Patients older than 70 years had a higher persistence than patients younger than 40 (HR 0.96 [0.53-0.9], Supplementary Figure 5A); concomitant csDMARDs and previous glucocorticoids were also associated with higher persistence in multivariable models, while sex and concomitant glucocorticoids were no longer associated (Supplementary Figure 5A).

**Figure 4.**
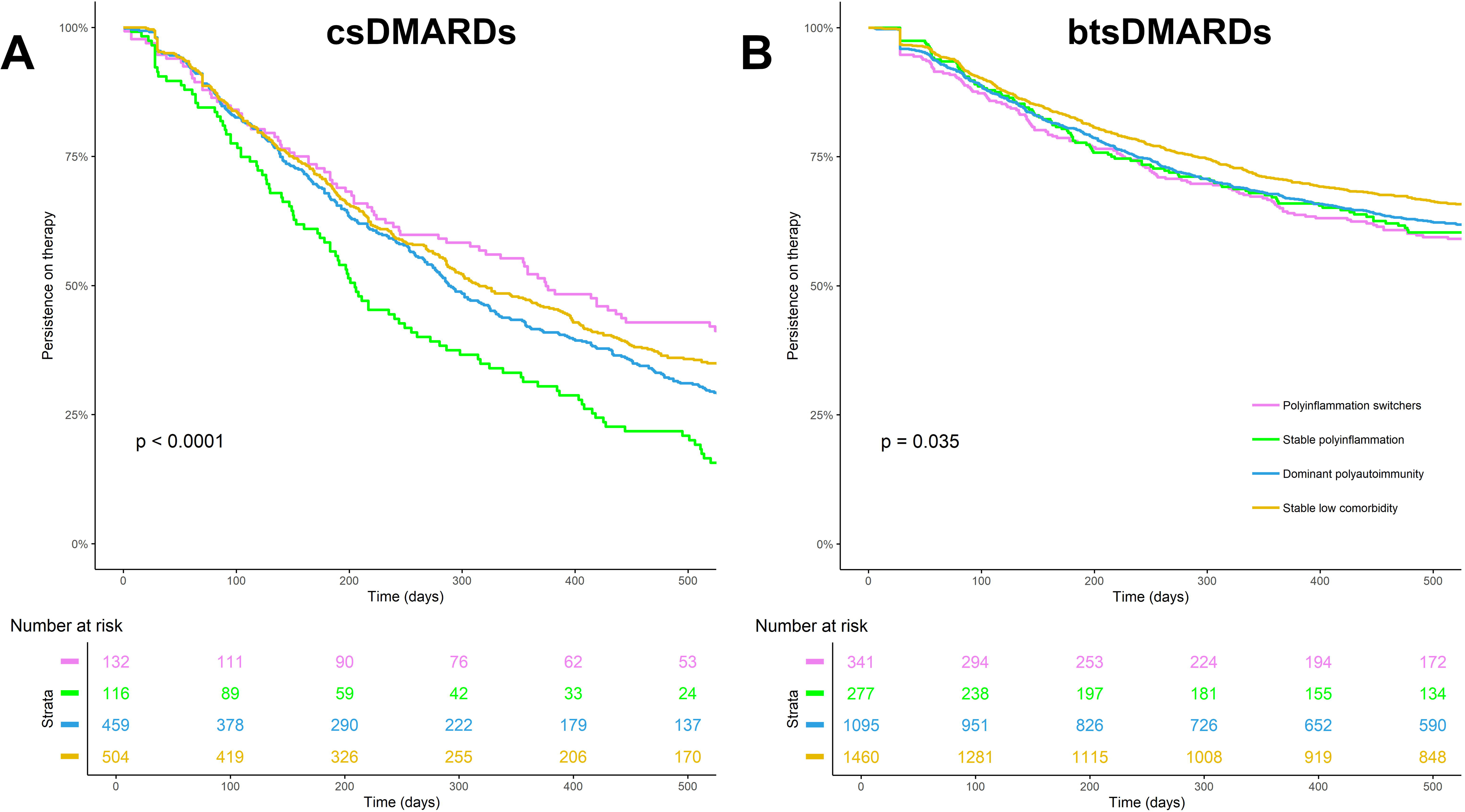
Kaplan-Meier of DMARD drug survival by comorbidity trajectory. (A) csDMARDs; (B) btsDMARDs.

btsDMARDs have different mechanisms of action, and since the comorbidity profiles might represent different etiopathogenic groups there might be more important differences in persistence for different btsDMARD classes. We therefore compared persistence among btsDMARD classes separately for the 4 comorbidity trajectories. Only stable low comorbidity and polyinflammation switchers (Figure 5A, 5B) but not stable polyinflammation nor dominant polyautoimmunity had significant differences in persistence among btsDMARD classes (Figure 5C, 5D). A higher persistence of JAKi and TNFi compared to anti-IL6R in patients with stable low comorbidity was confirmed in multivariable Cox models (HR 0.69 [0.5; 0.96] and 0.75 [0.59; 0.96] respectively, Supplementary Figure 5B), while in polyinflammation switchers CTLA4-Fc, JAKi and TNFi all had a higher persistence than anti-IL6R (HR 0.51 [0.29; 0.9], 0.55 [0.31; 1] and 0.52 [0.33; 0.82] respectively, Supplementary Figure 5C).

**Figure 5.**
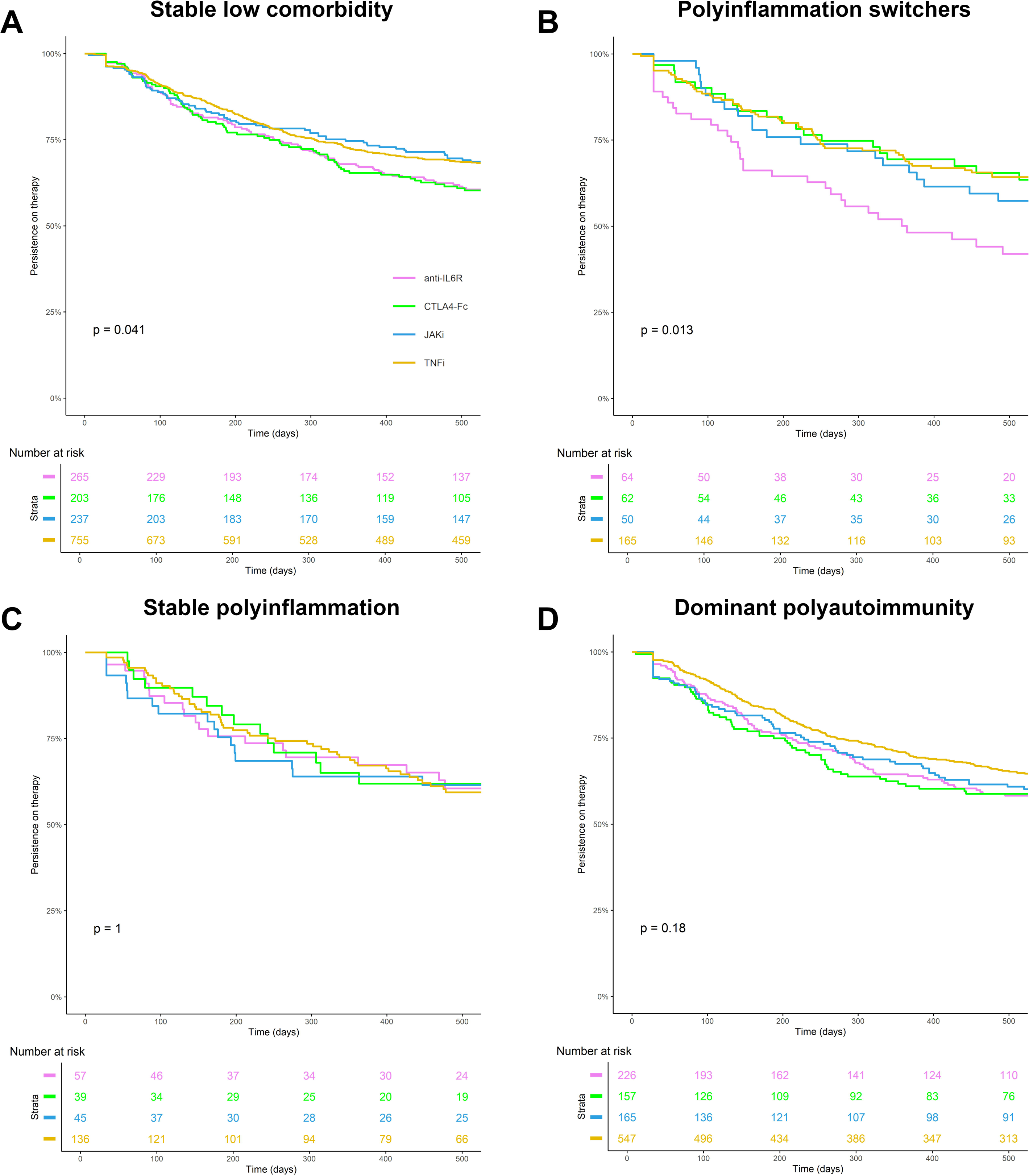
Kaplan-Meier of btsDMARD drug survival by therapeutic class. (A) stable low comorbidity; (B) Polyinflammation switchers; (C) Stable polyinflammation; (D) Dominant polyautoimmunity.

## Discussion

Heterogeneity in terms of disease activity, severity and response to therapy is well established in RA, yet studies to identify etiopathogenic clusters and their biological signatures are still at their infancy. In this work, we build on autoimmune and inflammatory comorbidity profiles previously identified as candidate etiopathogenic groups.^21^ We developed a model to classify RA patients into comorbidity profiles using 17 comorbidities; we characterized their temporal trajectories and investigated their usefulness as theranostic markers. We observed a relative stability of comorbidity patterns during 10 years and an increase in the proportion of patients with polyautoimmunity and polyinflammation during aging at a slow rate of 2-4% every decade. There were significant differences in persistence on DMARD therapy according to comorbidity trajectory, with the largest differences observed for csDMARDs where patients with stable polyinflammation had the lowest persistence. These results suggest that autoimmune and inflammatory comorbidity patterns might correspond to etiopathogenic groups and might be used for RA patient stratification in a therapeutic strategy perspective.

Since this study was performed on a retail-pharmacy drug dispensation database, we used algorithms to identify RA patients and comorbidities. RA patients selected by the BTSD algorithm must have taken at least one RA-indicated btsDMARD, meaning that patients who never switch from csDMARDs to btsDMARDs were excluded from the study. For the comorbidities, whenever available we used algorithms from the French Health Insurance Agency,^30^ yielding a similar prevalence in RA patients for anxiety, depression, hypertension and respiratory tract disorders, but a two-fold higher prevalence in LRx than in the French Health Insurance data^32^ for dyslipidemia. For the other comorbidities, we developed algorithms based on known drug indications. Similarly to what reported in the literature, 22% of RA patients had cardiovascular disorders,^33^ while cancer was less prevalent (2.7% in LRx vs 8.6% in the literature).^34^ Some differences were expected due to the more severe RA population selected by the BTSD algorithm and to the limited sensitivity in the detection of diseases such as cancer when using only pharmacy drug dispensations. However, our purpose was not to estimate the prevalence of comorbidities, but to classify patients in comorbidity profiles through a model using 17 comorbidities.

Although we expected that misclassifications in the comorbidities might have some repercussion on misclassification of the comorbidity profiles, we observed the same hierarchy of proportions of RA patients in the profiles in LRx as previously seen in TRANSIMMUNOM, albeit with more patients in the low comorbidity profile and less in polyautoimmunity and polyinflammation in LRx. This suggests that the model is robust and that most misclassifications are due to missed detection of polyautoimmunity or polyinflammation wrongly attributed to low comorbidity. Age and sex were also similarly distributed across the profiles in LRx and TRANSIMMUNOM RA patients.

Consistently with other reports in the literature for multimorbid RA patients, we observed a decreased usage of btsDMARDs in the polyautoimmunity and polyinflammation clusters.^35^ Although there were more patients in the polyautoimmunity and polyinflammation clusters taking no DMARDs, the intravenous bDMARDs are only delivered in hospitals and are not recorded in LRx, therefore this result might conceal an increased usage of intravenous bDMARDs in the multimorbid profiles. More RA patients in the polyautoimmune and polyinflammatory profiles used influenza vaccines, oral broad spectrum penicillins, antiseptics and disinfectants, suggesting that the multimorbid profiles have an increased risk of infections as previously reported.^36^

Previous studies have investigated the trajectory of the presence and burden of multimorbidity in RA. They followed the development of long-term conditions after RA diagnosis and they found a higher rate of comorbidity accrual in RA patients compared to non-RA controls.^4,37^ In this work, we report the first description of the temporal evolution of autoimmune and inflammatory comorbidity profiles. Due to regulatory constraints, we followed the patients for 10 years from entry in the database and we found 4 main trajectories, 3 of which characterized by the dominance of one of the 3 comorbidity patterns and one by a progressive switch from low comorbidity to polyinflammation. The limited switch from low comorbidity to multimorbid profiles during 10 years might be explained by the slow rate at which the proportion of patients in multimorbid profiles increased with aging. Interestingly the switch to polyautoimmunity started at a younger age than the switch to polyinflammation, and the rate of switch was significantly higher in RA than in non-regular controls comparable to the general population, as previously reported by others.^4,37^ 47% of RA patients older than 80 years were still in the low comorbidity cluster, suggesting that some patients never switch to the multimorbid profiles. We could not clearly isolate a trajectory with switches between polyautoimmunity and polyinflammation, meaning that 10 years follow-up is not sufficient to observe it, or that this trajectory is uncommon. It is therefore likely that polyautoimmunity and polyinflammation are two distinct multimorbid patterns reflecting different etiopathogenic entities. However, a limit of this analysis is that we did not have the date of diagnosis and the analyzed patients were a mixture of prevalent and incident cases, so we do not know the proportion of patients who are in multimorbid profiles already at RA onset. If certain switches happen early after diagnosis we may have missed them for the patients who entered in the database late after diagnosis.

This is the first study that compares therapy retention of several DMARDs among different comorbidity profiles in RA: others have studied the association of comorbidity indexes or of single comorbidities with efficacy of specific bDMARDs, and only a few clinical trials have compared safety and efficacy of different btsDMARDs according to patient multimorbidity.^11,12,14,38–40^ While a lower efficacy of DMARDs in multimorbid patients was known,^11^ here we describe a larger effect size of multimorbid trajectories on csDMARD than on btsDMARD retention. Patients with stable polyinflammation had the lowest retention on csDMARDs, suggesting that these patients might benefit from being started directly on btsDMARDs as first-line treatments.

Consistently with previous studies, concomitant csDMARDs were associated with an increased btsDMARD persistence.^41^ When comparing retention of different btsDMARDs separately by comorbidity trajectory we saw significant differences only in stable low comorbidity and polyinflammation switchers, with lower persistence of anti-IL6R in both trajectories and of CTLA4-Fc in stable low comorbidity only. Although we included in the analysis only the first initiation of btsDMARDs in the 10 years of follow-up, we could not determine the line of treatment, which might influence these differences. It is interesting to observe that polyinflammation switcher and stable polyinflammation trajectories did not necessarily follow the same patterns of association with DMARD persistence even after adjustment on age, suggesting that the impact of inflammation at early and late stages might be different.

Altogether, the results of the present work suggest that autoimmune and inflammatory comorbidity patterns might reflect etiopathogenic groups in RA. For the purpose of patient stratification, comorbidity profiles might be more appropriate than multimorbidity, which is more suitable for holistic care and prognosis. Until present, only a few clinical trials have investigated the usefulness of biomarkers such as synovial fluid B cell richness for RA patient stratification with contradictory results.^42,43^ While this research field is rapidly expanding^44^ and future investigations hold the promise to identify theranostic biomarkers, comorbidity profiles might be a simple tool for a first screening as only 17 comorbidities are needed for their classification. Once identified, biomarkers will allow a finer division relying on the main molecular actors of etiopathogenesis, but comorbidity profiles might still be a valuable complement by bringing information on the dominant autoimmune or inflammatory context in which these mechanisms take place.

As already mentioned this study has some limits. Comorbidities were derived from algorithms based on drug dispensations, yielding potential misclassifications. Information on several confounders such as disease duration, disease activity and severity, ACPA and RF seropositivity, line of DMARD treatment were not available, increasing the risk of unmeasured confounding in our analyses. The respective contribution of drug tolerance and therapy efficacy to DMARD retention could not be distinguished. The analyzed RA population is biased to more severe patients who switch to btsDMARDs and intravenous bDMARDs such as anti-CD20 could not be evaluated.

Strengths of this study are the high number of RA patients eligible for the analyses and the comparison with large healthy and non-RA control populations. The multinomial regression model developed based on 17 comorbidity variables may easily be applied by other researchers and clinicians to evaluate comorbidity profiles and is simpler than most comorbidity indexes based on more than 40 diseases.

In conclusion, autoimmune and inflammatory comorbidity profiles in RA are stable with a slow rate of switch from low comorbidity to either polyautoimmunity or polyinflammation, suggesting that they represent distinct etiopathogenic entities. DMARD retention is significantly different among comorbidity trajectories with particularly important size effect on first csDMARD initiation, highlighting the theranostic potential of comorbidity profiles in RA. These promising results need to be confirmed in a study on a larger RA population including patients who do not switch to btsDMARDs and they need to be extended to intravenous bDMARDs that were not evaluated in the present work.

## Data Availability

All data produced in the present study are available upon reasonable request to the authors

## Acknowledgments

This work was a collaboration between AP-HP and IQVIA. We thank all the epidemiologists and data scientists at IQVIA who have followed and given feedback to this work: Isabelle Bardoulat, Cedric Collin, Jérémie Pinoteau, Angéline Fabre, Eric Martinho, Alban Duruisseau, Raphaëlle Jallat and Clément Chastagnol. We thank Dr. El Hadi Zerdazi for discussions and feedback.

## Notes

This study was supported by Institut Carnot@ AP-HP, by IQVIA and a Laboratory of Excellence grant (n⍰ANR-11-IDEX-0004-02).

Conflicts of interest: Signe Hässler, Julie Aste, Francis Berenbaum, Michelle Rosenzwajg, David Klatzmann and Milka Maravic declare no conflicts of interest. Jérémie Sellam declares consultancies, honoraria, advisory board, and speakers’ fees from Biogen, Celltrion, Pfizer, BMS, MSD, Abbvie, Lilly, IBSA, Janssen, Novartis, Fresenius Kabi.

### Competing Interest Statement

Signe Hassler, Julie Aste, Francis Berenbaum, Michelle Rosenzwajg, David Klatzmann and Milka Maravic declare no conflicts of interest.
Jeremie Sellam declares consultancies, honoraria, advisory board, and speakers fees from Biogen, Celltrion, Pfizer, BMS, MSD, Abbvie, Lilly, IBSA, Janssen, Novartis, Fresenius Kabi.

### Funding Statement

This study was supported by Institut Carnot@ AP-HP, by IQVIA and a Laboratory of Excellence grant (n◦ANR-11-IDEX-0004-02).

### Author Declarations

The TRANSIMMUNOM study was performed according to the principles of the Helsinki declaration, written informed consent was collected for each patient and the study was approved by the local ethics committee in Ile de France (CPP-IdFVI approval number 48-15). The LRx database was authorized by the French Data Protection Authority (CNIL) on the 21st of October 2011 [reference: DE-2011-097] and updated on July 2018 for compliance with the GDPR [reference: DE-2018-289].

