## Supplementary materials, figures and tables for "Autoimmune and inflammatory comorbidity patterns in rheumatoid arthritis: temporal trajectories and impact on persistence on DMARD therapy"

### Supplementary methods

#### Study population

LRx (Lifelink Treatment Dynamics) covers nearly 45% of French retail pharmacies and includes around 45 million patients covered by the French Health Insurance Funds or other insurance schemes. It is representative of the general population in terms of geographical distribution across metropolitan France and the age of the covered population. The available variables include patients' characteristics (unique identification number, year of birth, gender), prescribers' details (geographical area of practice, private or hospital practice, specialty), and treatments (prescription and dispensing dates, packaging, dosage, "code identifiant de présentation" [CIP, a unique code for treatment packaging], dispense volume).

The BTSD algorithm identifies patients who received an oral or subcutaneous targeted therapy (TT) for a chronic inflammatory disease. For RA, the prescriber is a rheumatologist, the TT indication is RA and the tracking baskets included: i) Immunosuppressants prescribed at any time (leflunomide, methotrexate without antipsoriatic topical treatments, emollients, topical tacrolimus or dermocorticoids, or historical delivery just before TT initiation of sulfasalazine or hydroxychloroquine). ii) Co-prescription at the initiation of TT of immunosuppressants (methotrexate or leflunomide with oral corticosteroids without antipsoriatic topical treatments, emollients, topical tacrolimus or dermocorticoids). The tracking baskets excluded immunosuppressants such as ciclosporin, cyclophosphamide, or other biotherapies such as anti-IL1 or belimumab not indicated for RA.

#### Study variables

We developed algorithms to define 17 comorbidity variables from drug deliveries. For 7 of the 17 comorbidity variables two alternative algorithms were built, one with more stringent criteria (e.g. a higher threshold of the minimum number of deliveries of a disease-specific drug), one with less stringent criteria. A detailed definition of the non-immunological comorbidity algorithms with the EphMRA drug classes used is given in Supplementary Table 1 while the threshold frequency of drug delivery used is given in Supplementary Table 2. Immunological disorders were classified as autoimmune or inflammatory diseases according to McGonagle and McDermott classification and as previously described.[1,2] In order to count the number of autoimmune diseases and the number of inflammatory diseases, 34 algorithms were built. For autoimmune and inflammatory diseases that are treated with specific drugs other than RA DMARDs, each diagnosis was defined for a total of 12 autoimmune and 7 inflammatory comorbidity variables. For drugs that may be used for several diseases, we defined 3 autoimmune disease-specific, 2 inflammatory disease-specific drug variables and 9 mixed specificity drug variables whenever a drug could be used for both autoimmune and inflammatory diseases. An autoimmunity score was attributed to the mixed specificity drug variables by counting the number of autoimmune diseases for which it is indicated divided by the total number of AID for which it is indicated, and similarly an inflammatory score was defined as the ratio of the number of inflammatory diseases to the total number of AIDs for which it is indicated. A detailed definition of the 34 non-immunological comorbidity algorithms with the EphMRA drug classes used is given in Supplementary Table 3 while the threshold frequency of drug delivery used is given in Supplementary Table 4**.** For each patient, the number of autoimmune diseases (including the RA diagnosis) was calculated as the sum of each of the autoimmune diagnoses, the autoimmune disease-specific drugs, and the autoimmunity score of the mixed specificity drugs; the number of inflammatory diseases was calculated as the sum of the inflammatory diagnoses, the inflammatory disease-specific drugs and the inflammatory score of the mixed specificity drugs. In the stringent definition of number of autoimmune or inflammatory diseases the mixed-specificity drugs were not used (Supplementary Table 4). As an example, a patient with RA (autoimmune) and myasthenia gravis (autoimmune), taking mycophenolate mofetil (autoimmune-specific) and diprosalic (mixed specificity with score 0.5 for autoimmunity and inflammation) will have 3.5 autoimmune diseases and 0.5 inflammatory diseases with the non-stringent definition and 3 autoimmune diseases and 0 inflammatory diseases with the stringent definition.

For 7 out of the 17 comorbidity variables a stringent and a non-stringent algorithm variant were tested, generating $2^7$ datasets of different combinations of the variants. To select an algorithm for each comorbidity, we performed a sensitivity analysis by applying the best performing classification model to all the 128 datasets and verifying the variability in the proportion of patients classified in the comorbidity clusters. One of the 32 scenarios equally close to the median was randomly selected to be used for the rest of the analyses: this model uses the non-stringent variants for atopy, GI disease and hypertension and the stringent variants for allergy, cardiovascular disease, autoimmune and inflammatory disease number.

**Classification model**

To classify RA patients and controls into comorbidity clusters we developed a classification model on the TRANSIMMUNOM study. The number of autoimmune and of inflammatory diseases were cut into multinomial categories as previously described.[1]

327 AID patients and 48 healthy controls from the TRANSIMMUNOM study were split into 75% training and 25% test datasets. Three models were developed on the training dataset, using the comorbidity cluster as outcome and the comorbidities as predictors: a multinomial logistic regression, an elastic-net penalized multinomial logistic regression and a gradient boosted tree. Model hyperparameters (lambda for elastic-net; shrinkage, number of trees, interaction depth and minimum number of observations per node for gradient boosted tree) were optimized through 10-fold internal cross-validation on the training dataset to minimize the misclassification error. The models with the optimal parameters were applied to the test dataset to assess model performance in terms of accuracy, F1, sensitivity, specificity, PPV and NPV. The best performing model in terms of accuracy was selected to predict the comorbidity clusters of the RA patients and controls from the LRx database once per year for each year of follow-up.

### Supplementary results

#### Comorbidity profile classification model and sensitivity analysis

Three models were developed on the TRANSIMMUNOM study to classify patients into comorbidity clusters using 17 comorbidity variables. They all had a very high accuracy, with the best performance for the multinomial logistic regression and the gradient boosted tree (96.81 [90.96; 99.34] % accuracy for both, Supplementary Figure 2). Since multinomial logistic regression is a simpler model and performed equally well to the gradient boosted tree it was selected to be applied to the RA patients in LRx to predict their comorbidity profiles. The model coefficients are available in Supplementary Table 6. The model predicted correctly all the patients in the polyautoimmunity profile and only made a few misclassification errors on the low comorbidity and polyinflammation clusters (Supplementary Figure 2, Supplementary Table 7). Recall, specificity and F1 score were better for the low comorbidity than the polyinflammation profile (Supplementary Table 7).

As mentioned in the methods, we developed algorithms to define 17 comorbidities from drug dispensations in LRx on the eligible RA patients (Supplementary Tables 1-4). For 7 comorbidities two alternative algorithms, one with stringent and one with non-stringent criteria were defined and a sensitivity analysis was performed on the 128 ($2^7$) datasets generated from the possible combinations of the comorbidity definition variants. The multinomial regression classification model was applied to the 128 datasets and the range of percentage of patients attributed to each comorbidity profile was examined. The variability was relatively low in all the clusters, with lower levels in the polyinflammation cluster (range 2.64%, Supplementary Figure 3), and higher in polyautoimmunity and low comorbidity (range 5.78% and 6.13% respectively, Supplementary Figure 3). Since the comorbidity definition variants did not have a major impact on the percentage of RA patients attributed to the comorbidity profiles (Supplementary Figure 3), the variants corresponding to a scenario closest to the theoretical median were selected to be used for the rest of the analyses.


**Supplementary figure 1-** Study design scheme. (A) Assessment of comorbidities, comorbidity profiles and trajectories. (B) Case-control study on the first year of follow- up. (C) Cohort study of persistence on DMARDs.

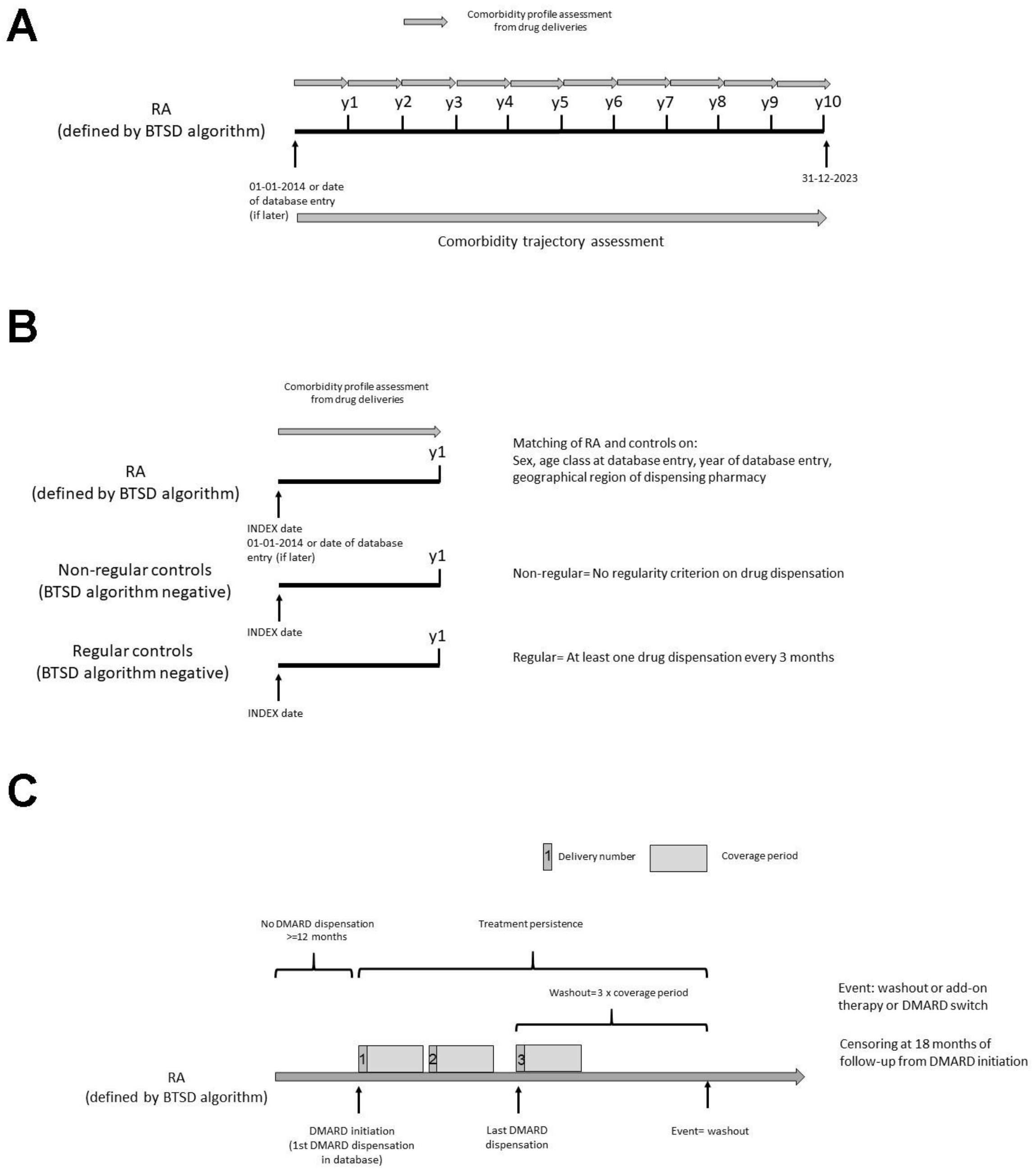


**Supplementary table 1-** EphMra codes used for non-immunological comorbidity definition (excel sheet)

**Supplementary table 2-** Algorithms used to define 15 non-immunological comorbidities.

| Comorbidity | Stringent algorithm | Non-stringent algorithm | Reference |
|---|---|---|---|
| Allergy | • At least 1 allergy drug<br>• At least 2 years of follow-up with allergy drug | • At least 2 allergy drugs<br>• At least 2 years of follow-up with allergy drug | |
| Atopy | At least 2 atopy drug deliveries | • At least 2 atopy drug deliveries<br>• At least 2 years of follow-up with atopy drug deliveries | |
| Cardiovascular disease | • At least 3 cardiovascular drug deliveries<br>• Beta-blockers excluded (C07A) | • At least 3 cardiovascular drug deliveries<br>• Beta-blockers included (C07A) | |
| Gastrointestinal disorder | • At least 2 GI disorder drug deliveries (Proton Pump Inhibitors excluded) OR at least 8 PPI deliveries.<br>• At least 2 years with these criteria | • At least 4 GI disorder drug deliveries<br>• At least 2 years with GI disorder drugs | |
| Hypertension | • At least 3 hypertension drug deliveries<br>• Beta-blockers excluded (C07A) | • At least 3 hypertension drug deliveries<br>• Beta-blockers included (C07A) | https://www.assurance-maladie.ameli.fr/sites/default/files/2024_methode-reperage-pathologies_cartographie.pdf |
| Anxiety | At least 3 anxyolitic drug deliveries | _ | https://www.assurance-maladie.ameli.fr/sites/default/files/2024_methode-reperage-pathologies_cartographie.pdf |
| Cancer | At least one cancer drug | _ | |
| Depression | At least 3 anti-depressant drug deliveries | _ | https://www.assurance-maladie.ameli.fr/sites/default/files/2024_methode-reperage-pathologies_cartographie.pdf |
| Dyslipidemia | At least 3 dyslipidemia drug deliveries | _ | https://www.assurance-maladie.ameli.fr/sites/default/files/2024_methode-reperage-pathologies_cartographie.pdf |
| Endocrine disorder | At least 3 endocrine disorder drug deliveries | _ | |
| Hepatobiliary disease | At least 2 hepatobiliary drug deliveries | _ | |
| Periodontal disease | • At least one antibiotic "bitherapy" of stomatological antibiotics (amoxicillin, clarithromycin, metronidazole) without PPI (<2 PPI/year) OR at least two deliveries of metronidazole and spiramycine OR at least 1 mouth wash/parodontal toothpaste product during the year | _ | |

| | | | |
|---|---|---|---|
| | • One of these conditions must be fulfilled during at least 2 years (not necessarily consecutive), otherwise abscess and wisdom tooth extraction are more likely<br>• Behcet patients excluded, i.e. patients taking mouth product AND a Behcet drug (apremilast, colchicine) | | |
| **Respiratory tract disorder** | • At least 3 R03 OR 4 R05 drug deliveries<br>• At least two years with R05 criteria, otherwise not chronic cough<br>• One single year with R03 criteria sufficient | _ | https://www.assurance-maladie.ameli.fr/sites/default/files/2024_methode-reperage-pathologies_cartographie.pdf |
| **Viral infection** | At least one antiviral drug | _ | |
| **Vision disorders** | At least 1 glaucoma or cataract drug OR 1 macular degeneration drug (if age >=80, excluding diabetes patients) | _ | |

**Supplementary table 3-** EphMra codes used for autoimmune and inflammatory comorbidity definition. (excel sheet)

**Supplementary table 4-** Algorithms used to define autoimmune and inflammatory comorbidities.

| Comorbidity/ drug variable | Stringent algorithm | Non-stringent algorithm | Autoimmunity score | Inflammation score |
|---|---|---|---|---|
| **Alopecia** | At least 1 alopecia drug | _ | 1 | 0 |
| **Hypothyroidism** | At least 2 hypothyroid drugs | _ | 1 | 0 |
| **Hyperthyroidism** | At least 1 hyperthyroidism drug | _ | 1 | 0 |
| **Immune thrombocytopenic purpura** | At least 1 ITP drug | _ | 1 | 0 |
| **Multiple Sclerosis** | At least 1 MS drug | _ | 1 | 0 |
| **Myasthenia Gravis** | At least 1 myasthenia drug AND no hypertension (Suppl. Table 2) | _ | 1 | 0 |
| **Narcolepsia** | At least 1 narcolepsia drug | _ | 1 | 0 |
| **Psoriasis** | At least 1 psoriasis drug OR 2 dermocorticoids (NOT LOCOID) AND no atopy (Suppl. Table 2) during the year | _ | 1 | 0 |
| **Raynaud's syndrome** | At least 1 Raynaud drug | _ | 1 | 0 |
| **Primary sclerosing cholangitis and biliary cirrhosis** | At least one biliary autoimmune disease drug | _ | 1 | 0 |

| | | | | |
|---|---|---|---|---|
| **Sjögren's syndrome** | At least 1 Sjögren drug AND no vision disorder (glaucoma, Suppl. Table 2) | _ | 1 | 0 |
| **Type 1 diabetes** | At least 3 T1D drug deliveries (any insulin) AND no oral or injectable antidiabetic | _ | 1 | 0 |
| **Rheumatoid arthritis (applied only to control populations)** | At least 1 RA csDMARD AND no cancer (Suppl. Table 2) | _ | 1 | 0 |
| **Behçet's disease** | At least 1 Behçet drug (apremilast or colchicine) AND one Behçet mouth wash | _ | 0 | 1 |
| **Genital lichen** | At least 1 genital lichen drug | _ | 0 | 1 |
| **Gout** | At least 1 gout drug AND no cancer (Suppl. Table 2) | _ | 0 | 1 |
| **Inflammatory Bowel Disease** | At least one IBD drug | _ | 0 | 1 |
| **Osteoarthritis** | At least 1 osteoarthritis drug | _ | 0 | 1 |
| **Type 2 diabetes** | At least 1 T2D drug AND no rapid insulin | _ | 0 | 1 |
| **Uveitis** | At least 1 uveitis drug AND no allergy (Suppl. Table 2) AND no vision disorder (glaucoma, cataract; Suppl. Table 2) | _ | 0 | 1 |
| **Mycophenolate or tacrolimus** | At least 1 mycophenolate or tacrolimus, excluding T2D (Suppl. Table 2) and uveitis | _ | 1 | 0 |
| **Disulone** | At least one disulone | _ | 1 | 0 |
| **Acitretin (AI)** | At least one acitretin AND no psoriasis | _ | 0.33 | 0.67 |
| **Apremilast (AI)** | At least one apremilast AND no psoriasis nor Behçet | _ | 0.5 | 0.5 |
| **Diprosalic (AI)** | At least one diprosalic AND no psoriasis | _ | 0.5 | 0.5 |
| **Ixekizumab (AI)** | At least one ixekizumab AND no psoriasis | _ | 0.5 | 0.5 |
| **Vitamin B12 (AI)** | At least one vitamin B12 AND no IBD (indicated for ileotomy for Crohn's) | _ | 0.5 | 0.5 |
| **Alitretinoin (AI)** | At least one alitretinoin AND no psoriasis nor atopic dermatitis (Suppl. Table 2) | _ | 0.67 | 0.33 |
| **Locoid (AI)** | At least one locoid excluding atopic dermatitis (Suppl. Table 2) and psoriasis | _ | 0.67 | 0.33 |
| **Nerisone (AI)** | At least one nerisone AND no psoriasis nor atopic dermatitis (Suppl. Table 2) | _ | 0.67 | 0.33 |
| **Methoxsalen (AI)** | At least one methoxsalen AND no psoriasis nor alopecia | _ | 0.75 | 0.25 |
| **Azathioprine (AI)** | At least one azathioprine AND no ITP, IBD nor myasthenia | _ | 0.89 | 0.11 |

| | | | | |
|---|---|---|---|---|
| **Colchicine** | At least one colchicine AND no Behçet nor gout | _ | 0 | 1 |
| **Iluvien** | At least one iluvien AND no T2D nor uveitis | _ | 0 | 1 |
| **Urarthone** | At least one urarthone AND no gout nor osteoarthritis | _ | 0 | 1 |
| **Number of autoimmune diseases** | Sum of the A score of AID comorbidities and drugs, AI drugs excluded | Sum of the A score of AID comorbidities and drugs, AI drugs included | _ | _ |
| **Number of inflammatory diseases** | Sum of the I score of AID comorbidities and drugs, AI drugs excluded | Sum of the I score of AID comorbidities and drugs, AI drugs included | _ | _ |

**Supplementary table 5-** EphMra codes used for DMARDs and oral glucocorticoids (excel sheet).

**Supplementary figure 2-** Confusion matrix of predicted versus true comorbidity clusters using three different classification models.

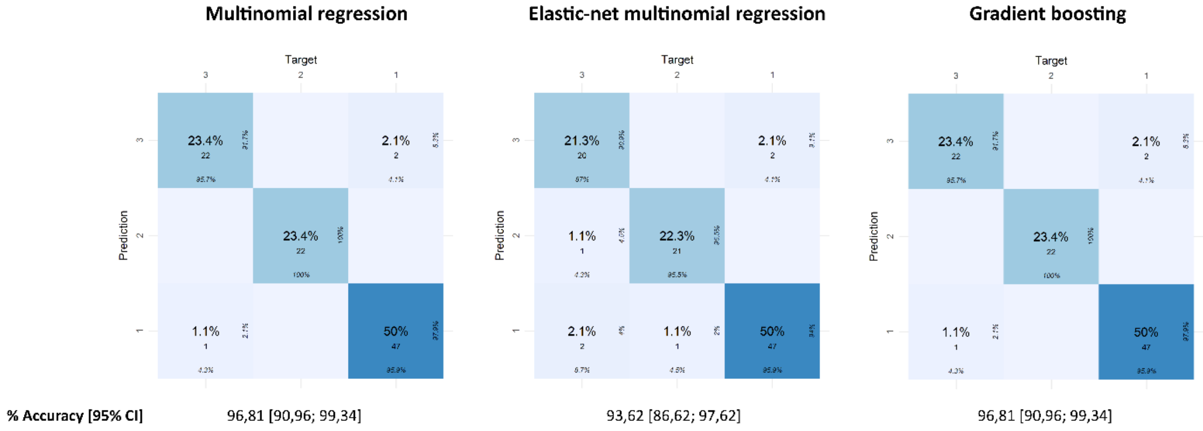


**Supplementary table 6-** Multinomial logistic regression classification model coefficients. The low comorbidity cluster is used as reference (excel sheet).

**Supplementary table 7-** Performance metrics, global and for each class - Multinomial logistic regression model. PPV- Positive Predictive Value; NPV- Negative Predictive Value

| Class | Recall (%) | Specificity (%) | PPV(%) | NPV (%) | F1 score (%) |
|---|---|---|---|---|---|
| Low comorbidity | 95,92 | 97,78 | 97,92 | 95,65 | 96,91 |
| Polyautoimmunity | 100 | 100 | 100 | 100 | 100 |
| Polyinflammation | 95,65 | 97,18 | 91,67 | 98,57 | 93,62 |
| Overall performance (all classes) | 97,2 | 98,3 | 96,5 | 98,1 | 96,8 |

**Supplementary figure 3-** Boxplot of percentage of RA patients in LRx attributed to comorbidity profiles on 128 datasets using different combinations of comorbidity definition variants.

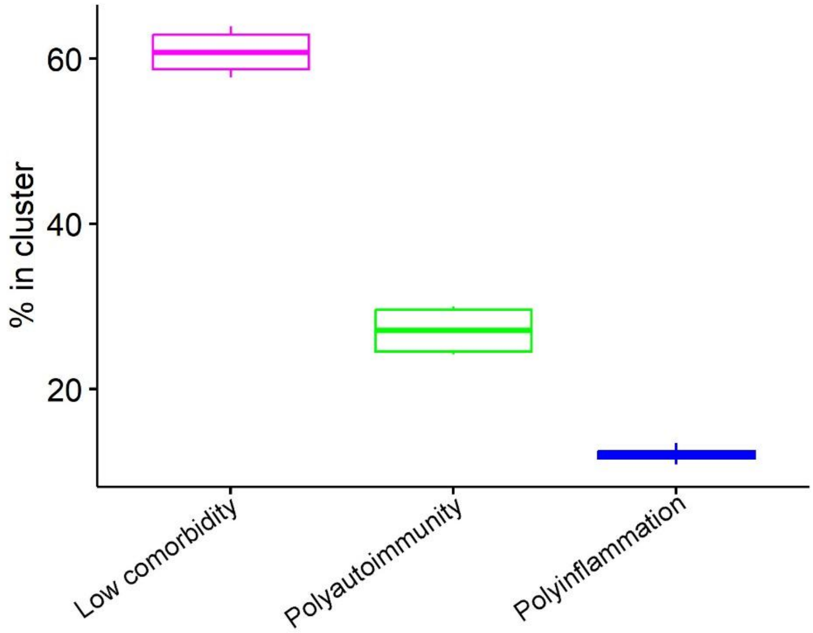


**Supplementary table 8-** Comparison of percentage of patients in comorbidity clusters and demographic characteristics in the clusters between RA patients of the TRANSIMMUNOM study and of the LRx database.

| | | Low comorbidity | Polyautoimmunity | Polyinflammation | p-value |
|---|---|---|---|---|---|
| **Comorbidity profile (N, %)** | | | | | |
| | TRANSIMMUNOM | 42 (53.2%) | 23 (29.1%) | 14 (17.7%) | - |
| | LRx | 9403 (61.9%) | 3745 (24.7%) | 2041 (13.4%) | - |
| **Age (years) Mean (SD)** | | | | | |
| | TRANSIMMUNOM | 47.81 (13.27) | 48.7 (15.35) | 56.57 (10.31) | 0.06 |
| | LRx | 55.9 (12) | 59.6 (11.7) | 62.1 (10.9) | <0.001 |
| **Sex (N Female, %)** | | | | | |
| | TRANSIMMUNOM | 32 (76.2%) | 21 (91.3 %) | 10 (71.4%) | 0.21 |
| | LRx | 5671 (60.6%) | 2807 (75.5%) | 1270 (62.5%) | <0.001 |

**Supplementary table 9-** Top 10 EphMra ATC 4 classes of concomitant medications most frequently taken by RA patients during the first year of follow-up in LRx. Comparison between comorbidity profiles.

| | Low comorbidity (N=9403) | Polyautoimmunity (N=3745) | Polyinflammation (N=2041) | p-value |
|---|---|---|---|---|
| **Non-narcotics and antipyretics** | | | | |
| Yes | 7246 (77.1%) | 3222 (86.0%) | 1806 (88.5%) | <0.001 |
| No | 2157 (22.9%) | 523 (14.0%) | 235 (11.5%) | |
| **Proton pump inhibitors** | | | | |
| Yes | 4827 (51.3%) | 2440 (65.2%) | 1623 (79.5%) | <0.001 |
| No | 4576 (48.7%) | 1305 (34.8%) | 418 (20.5%) | |
| **Other anti-anaemic products, including folic acid, folinic acid** | | | | |
| Yes | 4403 (46.8%) | 1654 (44.2%) | 890 (43.6%) | 0.00254 |
| No | 5000 (53.2%) | 2091 (55.8%) | 1151 (56.4%) | |
| **Vitamin D** | | | | |
| Yes | 3696 (39.3%) | 1909 (51.0%) | 1028 (50.4%) | <0.001 |
| No | 5707 (60.7%) | 1836 (49.0%) | 1013 (49.6%) | |
| **Anti-rheumatic, non-steroidal, plain** | | | | |
| Yes | 3990 (42.4%) | 1604 (42.8%) | 871 (42.7%) | 0.91 |
| No | 5413 (57.6%) | 2141 (57.2%) | 1170 (57.3%) | |
| **Influenza vaccines** | | | | |
| Yes | 2572 (27.4%) | 1514 (40.4%) | 946 (46.3%) | <0.001 |
| No | 6831 (72.6%) | 2231 (59.6%) | 1095 (53.7%) | |
| **Oral broad spectrum penicillins** | | | | |
| Yes | 2276 (24.2%) | 1203 (32.1%) | 688 (33.7%) | <0.001 |
| No | 7127 (75.8%) | 2542 (67.9%) | 1353 (66.3%) | |
| **Topical anti-rheumatics** | | | | |
| Yes | 1967 (20.9%) | 1056 (28.2%) | 602 (29.5%) | <0.001 |
| No | 7436 (79.1%) | 2689 (71.8%) | 1439 (70.5%) | |
| **Antiseptics and disinfectants** | | | | |
| Yes | 1668 (17.7%) | 949 (25.3%) | 562 (27.5%) | <0.001 |
| No | 7735 (82.3%) | 2796 (74.7%) | 1479 (72.5%) | |
| **Topical nasal corticosteroids without antibacterials** | | | | |
| Yes | 1425 (15.2%) | 1276 (34.1%) | 457 (22.4%) | <0.001 |
| No | 7978 (84.8%) | 2469 (65.9%) | 1584 (77.6%) | |

**Supplementary figure 4-** Transversal frequency of states (left panels) and mean time spent in each state (right panels) of identified clusters of sequences with similar temporal trajectory.

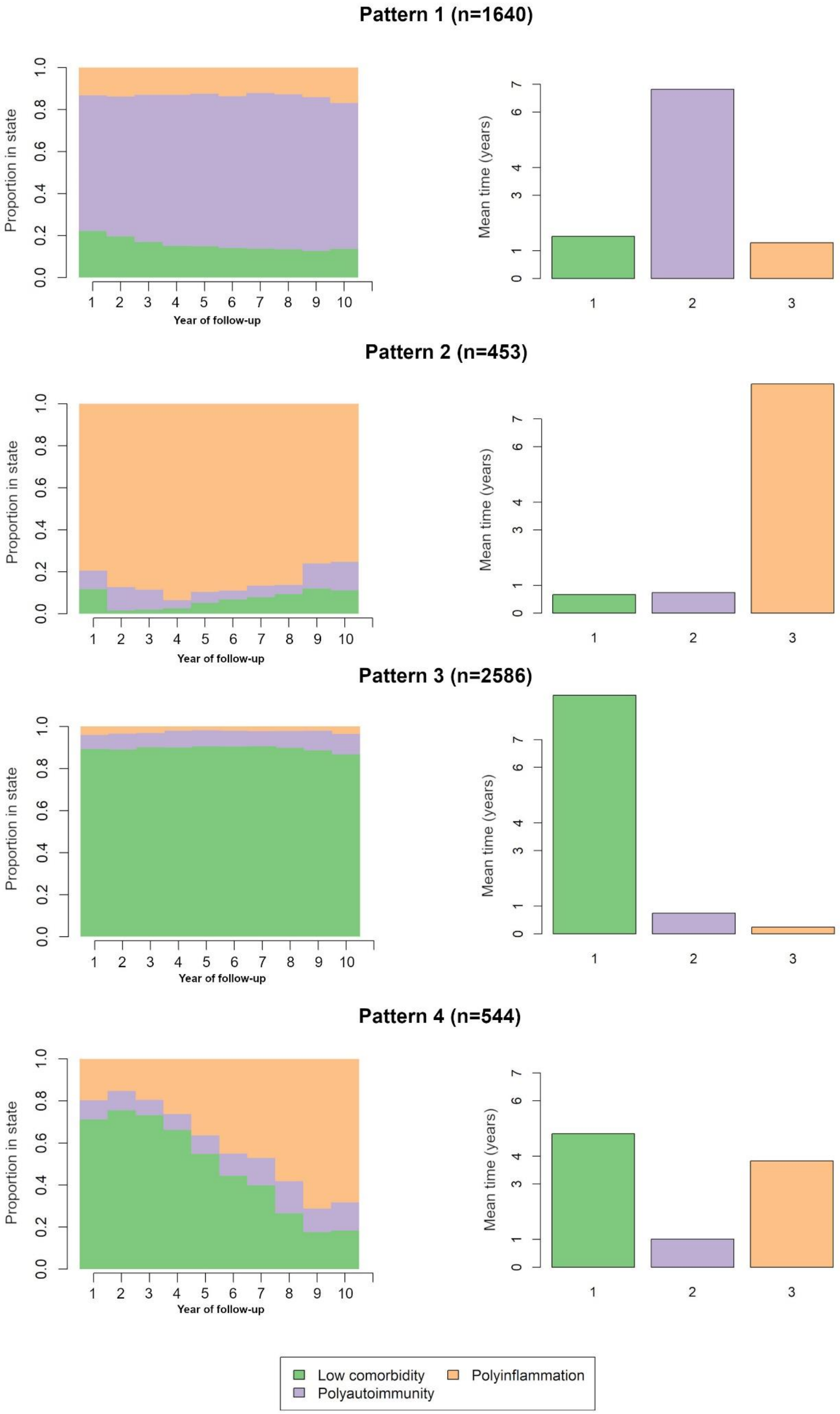

Pattern 1 (n=1640)
Pattern 2 (n=453)
Pattern 3 (n=2586)
Pattern 4 (n=544)
Proportion in state
Year of follow-up
Mean time (years)
Low comorbidity
Polyautoimmunity
Polyinflammation

**Supplementary table 10-** Linear regression and correlation coefficients of proportion of patients in comorbidity clusters with 5-year ordered age classes. RA: Rheumatoid arthritis; CI: Confidence Interval; rho: Pearson's correlation coefficient

| Population | Comorbidity profile | β | 95% CI of β | rho | Correlation p-value |
|---|---|---|---|---|---|
| RA | Low comorbidity | -0.0063 | [-0.0073 ; -0.0053] | -0.967 | $<10^{-8}$ |
| Regular controls | Low comorbidity | -0.0064 | [-0.0070 ; -0.0057] | -0.987 | $<10^{-10}$ |
| Non-regular controls | Low comorbidity | -0.0037 | [-0.0043 ; -0.003] | -0.964 | $<10^{-8}$ |
| RA | Polyautoimmunity | 0.0025 | [0.0009 ; 0.0041] | 0.675 | 0.0058 |
| Regular controls | Polyautoimmunity | 0.00039 | [-0.00012 ; 0.0009] | 0.416 | 0.12 |
| Non-regular controls | Polyautoimmunity | 0.00076 | [0.00057 ; 0.00095] | 0.925 | $<10^{-6}$ |
| RA | Polyinflammation | 0.0038 | [0.0036 ; 0.0047] | 0.938 | $<10^{-6}$ |
| Regular controls | Polyinflammation | 0.006 | [0.0056 ; 0.0064] | 0.994 | $<10^{-13}$ |
| Non-regular controls | Polyinflammation | 0.0029 | [0.0024 ; 0.0034] | 0.962 | $<10^{-7}$ |

**Supplementary table 11-** Demographic characteristics and comorbidity profiles of RA-matched control populations in LRx.

| | RA (N=15189) | Regular controls (N=30188) | Non-regular controls (N=30121) |
|---|---|---|---|
| **Age (years)** | | | |
| Mean (SD) | 57.6 (12.0) | 58.9 (13.7) | 57.1 (14.2) |
| Median [Min, Max] | 58.0 [19.0, 97.0] | 60.0 [19.0, 97.0] | 57.0 [19.0, 97.0] |
| **Sex** | | | |
| Male | 5359 (35.3%) | 10712 (35.5%) | 10690 (35.5%) |
| Female | 9748 (64.2%) | 19476 (64.5%) | 19431 (64.5%) |
| Missing | 82 (0.5%) | 0 (0%) | 0 (0%) |
| **Year of start follow-up** | | | |
| 2014 | 4804 (31.6%) | 9535 (31.6%) | 9332 (31.0%) |
| 2015 | 398 (2.6%) | 771 (2.6%) | 878 (2.9%) |
| 2016 | 405 (2.7%) | 794 (2.6%) | 824 (2.7%) |
| 2017 | 1089 (7.2%) | 2168 (7.2%) | 2190 (7.3%) |
| 2018 | 1689 (11.1%) | 3357 (11.1%) | 3373 (11.2%) |
| 2019 | 1980 (13.0%) | 3947 (13.1%) | 3944 (13.1%) |
| 2020 | 3250 (21.4%) | 6472 (21.4%) | 6455 (21.4%) |
| 2021 | 1557 (10.3%) | 3144 (10.4%) | 3122 (10.4%) |
| 2022 | 17 (0.1%) | 0 (0%) | 3 (0.0%) |
| **Comorbidity cluster** | | | |
| Low comorbidity | 9403 (61.9%) | 17830 (59.1%) | 26580 (88.2%) |
| Polyautoimmunity | 3745 (24.7%) | 5042 (16.7%) | 1615 (5.4%) |
| Polyinflammation | 2041 (13.4%) | 7316 (24.2%) | 1926 (6.4%) |

**Supplementary figure 5-** Multivariate Cox proportional hazard models of persistence on btsDMARDs. (A) All btsDMARDs. (B) btsDMARDs by therapeutic class, stable low comorbidity patients only. (C) btsDMARDs by therapeutic class, polyinflammation switchers only.

## A

#### All btsDMARDs

| Variable | HR (95%CI) | P value |
|---|---|---|
| No. of patients | 3140 | |
| Comorbidity trajectory | | |
| Stable low comorbidity | 1.00 ( Reference ) | |
| Dominant polyautoimmunity | 1.19 ( 1.04 - 1.37 ) | 0.013 |
| Stable polyinflammation | 1.27 ( 1.02 - 1.58 ) | 0.032 |
| Polyinflammation switchers | 1.32 ( 1.09 - 1.6 ) | 0.005 |
| Sex | | |
| Male | 1.00 ( Reference ) | |
| Female | 1.12 ( 0.98 - 1.28 ) | 0.104 |
| Age | | |
| <40 | 1.00 ( Reference ) | |
| [40,50) | 0.98 ( 0.75 - 1.27 ) | 0.86 |
| [50,70) | 0.87 ( 0.69 - 1.1 ) | 0.24 |
| >=70 | 0.69 ( 0.53 - 0.9 ) | 0.006 |
| Concomitant csDMARD | | |
| No | 1.00 ( Reference ) | |
| Yes | 0.86 ( 0.76 - 0.97 ) | 0.014 |
| Concomitant glucocorticoid | | |
| No | 1.00 ( Reference ) | |
| Yes | 0.98 ( 0.86 - 1.11 ) | 0.74 |
| Previous glucocorticoid | | |
| No | 1.00 ( Reference ) | |
| Yes | 0.69 ( 0.52 - 0.91 ) | 0.009 |

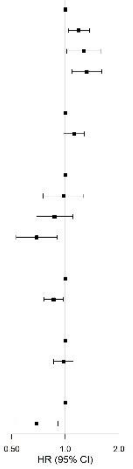


## B

#### btsDMARDs by therapeutic class, stable low comorbidity patients

| Variable | HR (95%CI) | P value |
|---|---|---|
| No. of patients | 1446 | |
| btsDMARD class | | |
| Anti-IL6R | 1.00 ( Reference ) | |
| CTLA4-Fc | 1.10 ( 0.81 - 1.49 ) | 0.54 |
| JAKi | 0.69 ( 0.5 - 0.96 ) | 0.027 |
| TNFi | 0.75 ( 0.59 - 0.96 ) | 0.02 |
| Sex | | |
| Male | 1.00 ( Reference ) | |
| Female | 0.99 ( 0.81 - 1.21 ) | 0.93 |
| Age | | |
| <40 | 1.00 ( Reference ) | |
| [40,50) | 1.07 ( 0.76 - 1.5 ) | 0.7 |
| [50,70) | 0.83 ( 0.61 - 1.14 ) | 0.25 |
| >=70 | 0.66 ( 0.44 - 0.97 ) | 0.03 |
| Concomitant csDMARD | | |
| No | 1.00 ( Reference ) | |
| Yes | 0.82 ( 0.68 - 1 ) | 0.04 |
| Concomitant glucocorticoid | | |
| No | 1.00 ( Reference ) | |
| Yes | 0.94 ( 0.76 - 1.16 ) | 0.56 |
| Previous glucocorticoid | | |
| No | 1.00 ( Reference ) | |
| Yes | 0.69 ( 0.47 - 1.01 ) | 0.055 |

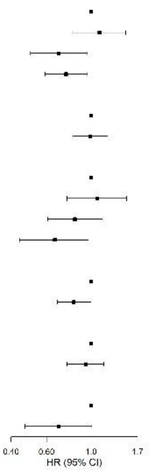


## C

#### btsDMARDs by therapeutic class, polyinflammation switchers

| Variable | HR (95%CI) | P value |
|---|---|---|
| No. of patients | 339 | |
| btsDMARD class | | |
| Anti-IL6R | 1.00 ( Reference ) | |
| CTLA4-Fc | 0.51 ( 0.29 - 0.9 ) | 0.02 |
| JAKi | 0.55 ( 0.31 - 1 ) | 0.052 |
| TNFi | 0.52 ( 0.33 - 0.82 ) | 0.005 |
| Sex | | |
| Male | 1.00 ( Reference ) | |
| Female | 0.87 ( 0.6 - 1.25 ) | 0.46 |
| Age | | |
| <40 | 1.00 ( Reference ) | |
| [40,50) | 0.69 ( 0.19 - 2.46 ) | 0.57 |
| [50,70) | 0.85 ( 0.26 - 2.79 ) | 0.78 |
| >=70 | 0.84 ( 0.25 - 2.83 ) | 0.78 |
| Concomitant csDMARD | | |
| No | 1.00 ( Reference ) | |
| Yes | 0.89 ( 0.63 - 1.27 ) | 0.53 |
| Concomitant glucocorticoid | | |
| No | 1.00 ( Reference ) | |
| Yes | 0.85 ( 0.58 - 1.24 ) | 0.41 |
| Previous glucocorticoid | | |
| No | 1.00 ( Reference ) | |
| Yes | 0.52 ( 0.16 - 1.67 ) | 0.27 |

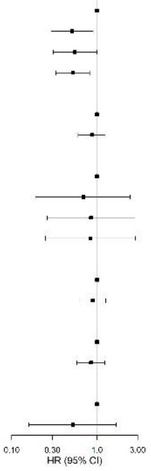
